# Mifepristone Priming with Misoprostol versus Intracervical Foley’s Catheter with Misoprostol for Induction of Labour in Late Second and Third Trimester Intrauterine Fetal Death: A Prospective Comparative Study

**DOI:** 10.64898/2026.08.06.26359872

**Authors:** Babita Das, Pratibha Garg

## Abstract

**Introduction:** Intrauterine fetal death (IUFD) beyond 24 weeks of gestation, particularly when accompanied by an unfavourable cervix, poses a distinct obstetric challenge in achieving safe and timely vaginal delivery while minimising maternal distress. Mifepristone priming followed by misoprostol and intracervical Foley’s catheter combined with misoprostol are both established approaches for cervical ripening and induction of labour in this setting, but direct comparative data especially from Indian tertiary care populations remain limited.

**Methods:** This prospective comparative study was conducted in the Department of Obstetrics and Gynaecology, Kamla Raja Hospital, Gajra Raja Medical College (GRMC), Gwalior, Madhya Pradesh, India, over a two-year period (November 2020–October 2022). One hundred and fourteen women with ultrasonography-confirmed IUFD beyond 24 weeks of gestation were alternately allocated to Group A (n=57; oral mifepristone 200 mg followed by gestational-age-adjusted vaginal misoprostol) or Group B (n=57; intracervical 16F Foley’s catheter followed by gestational-age-adjusted vaginal misoprostol). Outcomes assessed included pre- and post-induction Bishop score, induction-to-delivery interval, misoprostol dose requirement, need for oxytocin augmentation, mode of delivery, blood loss, maternal complications, pain (visual analogue scale, VAS), and patient satisfaction.

**Results:** Baseline age, parity, gestational age, and pre-induction Bishop score were comparable between groups (p>0.05). The mean post-induction (24-hour) Bishop score was significantly higher in Group A (7.39±2.07) than Group B (6.37±1.89; p=0.007). The mean induction-to-delivery interval was significantly shorter in Group A (25.43±6.84 hours) than Group B (29.26±5.54 hours; p=0.0014), and the median misoprostol dose requirement was significantly lower in Group A (50 mcg) than Group B (100 mcg; p<0.01). Mode of delivery, blood loss, oxytocin augmentation requirement, and overall maternal complication rates did not differ significantly between groups (all p>0.05). Pain scores were significantly lower in Group A (VAS 2.83±1.16) than Group B (VAS 6.18±1.69; p<0.0001), while patient satisfaction was comparable between groups (96.5% vs. 91.23%; p=0.244).

**Conclusions:** Both mifepristone-misoprostol and Foley’s catheter-misoprostol regimens are safe and effective methods for induction of labour following IUFD beyond 24 weeks of gestation with an unfavourable cervix. Mifepristone priming achieved a shorter induction-to-delivery interval, lower total misoprostol requirement, and substantially less procedural pain, making it an attractive first-line option where available, while Foley’s catheter remains a safe, low-cost, and widely accessible alternative, notwithstanding lower patient comfort.

## Introduction

Intrauterine fetal death (IUFD), or stillbirth, complicates a substantial proportion of pregnancies worldwide and represents one of the most emotionally and clinically demanding scenarios in obstetric practice. When IUFD is diagnosed beyond 24 weeks of gestation, timely induction of labour is generally recommended over expectant management, both to reduce maternal risks of coagulopathy and infection and to shorten the psychological burden of carrying a deceased fetus. However, many such pregnancies present with an unfavourable (unripe) cervix, particularly at earlier gestational ages, making cervical ripening a prerequisite for successful vaginal delivery.

Several pharmacological and mechanical methods are available for cervical ripening and induction of labour in this setting. Misoprostol, a synthetic prostaglandin E1 analogue, is widely used owing to its low cost, stability at room temperature, and multiple routes of administration. Mifepristone, an antiprogestin, sensitises the myometrium and cervix to the effects of prostaglandins through progesterone-receptor antagonism, and has been shown in several studies to shorten the induction-to-delivery interval when used as a priming agent prior to misoprostol. ^[1,2]^ Mechanical methods, particularly the intracervical Foley’s balloon catheter, achieve cervical ripening through direct mechanical dilatation and local release of endogenous prostaglandins, and are valued for their low cost, wide availability, and favourable safety profile, including in women with a prior caesarean scar. ^[3]^

Randomised and observational comparisons of mifepristone-misoprostol against Foley’s catheter-misoprostol regimens have been reported from varied obstetric populations, including term and post-term pregnancies and IUFD, with generally consistent findings of a shorter induction-to-delivery interval and lower analgesic requirement with mifepristone priming, though patient populations, gestational age ranges, and dosing protocols vary considerably between studies. ^[1,4,5]^ Comparative data specific to IUFD beyond 24 weeks of gestation, and specific to Indian tertiary-care obstetric populations, remain comparatively limited, which was the impetus for the present study.

### Aims and objectives

The aim of this study was to compare two regimens for induction of labour in late second and third trimester IUFD (gestational age >24 weeks): (i) mifepristone priming with misoprostol and (ii) intracervical Foley’s catheter with misoprostol. The specific objectives were to compare the two regimens with respect to induction-to-delivery time interval, mean misoprostol dose requirement, need for oxytocin augmentation, procedural pain, and patient satisfaction.

## Materials and Methods

### Study design and setting

This was a prospective comparative study conducted in the Department of Obstetrics and Gynaecology, Kamla Raja Hospital, Gajra Raja Medical College (GRMC), Gwalior, Madhya Pradesh, India, over a two-year period from November 2020 to October 2022.

### Sample size and allocation

A total of 114 women with IUFD were enrolled and alternately allocated into two groups of 57 each: Group A received oral mifepristone with misoprostol, and Group B received an intracervical Foley’s catheter with misoprostol.

### Eligibility criteria

#### Inclusion criteria

Pregnant women admitted with intrauterine fetal death diagnosed by real-time ultrasonography, with gestational age >24 weeks.

#### Exclusion criteria

Refusal of consent; gestational age <24 weeks; multiple pregnancy; eclampsia; cephalopelvic disproportion; malpresentation (transverse or oblique lie); previous two caesarean sections beyond 34 weeks; previous more than three caesarean sections; active lower genital tract infection; and deranged coagulation profile.

### Intervention protocols

After informed consent, a detailed history (age, parity, socioeconomic status, antenatal care, and relevant medical/obstetric history) was recorded, gestation confirmed by last menstrual period or earliest ultrasound, and a general, systemic, and obstetric examination performed, including per-speculum and per-vaginal assessment of the modified Bishop score. Baseline investigations and a coagulation profile were obtained in all participants.

#### Group A (n=57)

Tablet mifepristone 200 mg orally at admission. Bishop score was reassessed after 24 hours; if <6, gestational-age-adjusted vaginal misoprostol was administered in the posterior fornix every 4 hours (maximum 4 doses) until active labour was established. Failure to achieve a favourable cervix after 4 doses was classified as failed induction.

#### Group B (n=57)

A 16F Foley’s catheter was placed through the internal cervical os under aseptic conditions and inflated with 30 mL of sterile saline. After 12–24 hours, or spontaneous expulsion of the balloon, the Bishop score was reassessed; if <6, gestational-age-adjusted vaginal misoprostol was administered every 4 hours (maximum 4 doses) in the same manner as Group A.

**Table 1.** Gestational-age-adjusted first vaginal misoprostol dose.

| Gestational age | Misoprostol 1st vaginal dose |
| --- | --- |
| 24–30 weeks | 200 mcg |
| 31–35 weeks | 100 mcg |
| 36–41 weeks | 50 mcg |

Labour was augmented with an oxytocin infusion where clinically indicated. At 24 hours post-delivery, a structured patient satisfaction questionnaire assessed pain at catheter/tablet insertion, pain during cervical ripening (rated on a 0–10 visual analogue scale, where 0 = no pain and 10 = worst possible pain), analgesic use, and overall satisfaction with the induction process.

### Statistical analysis

Data were analysed using SPSS software. Continuous variables were expressed as mean ± standard deviation and compared using appropriate parametric tests; categorical variables were compared using the chi-square test. A p-value <0.05 was considered statistically significant.

**Figure 0.**
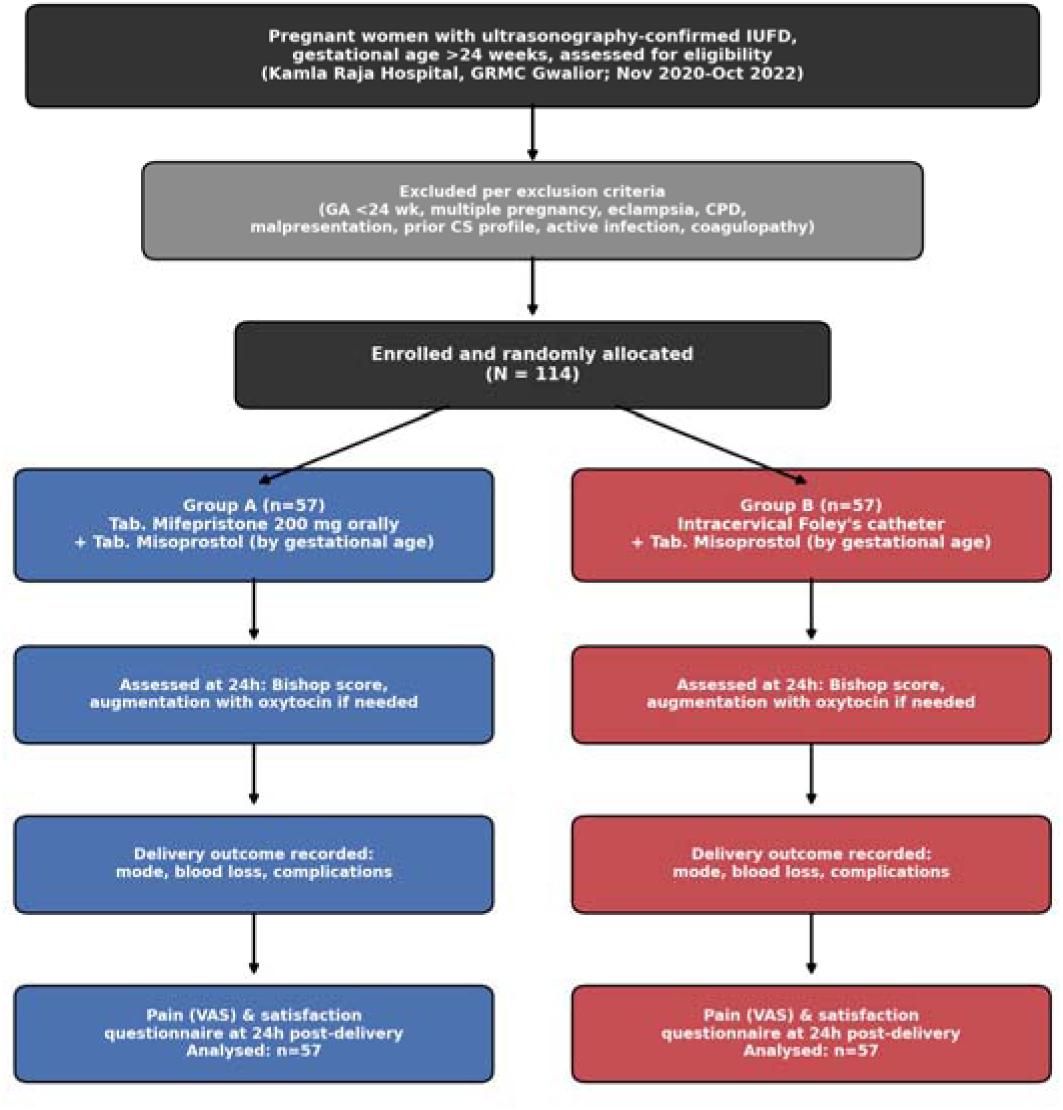
Study flow diagram summarising enrolment, allocation, intervention, and outcome assessment.

## Results

A total of 114 patients fulfilling the inclusion criteria were enrolled and divided equally into Group A (mifepristone + misoprostol, n=57) and Group B (Foley’s catheter + misoprostol, n=57).

### Baseline characteristics

**Table 2.** Age-wise distribution of the study population.

|  | Group A (n=57) | Group B (n=57) | p-value |
| --- | --- | --- | --- |
| Mean age ± SD (years) | 27.11 ± 2.99 | 26.39 ± 3.46 | 0.2379 |

The mean age of participants was comparable between the two groups (p=0.2379).

**Table 3.** Parity-wise distribution of the study population.

| Parity | Group A n (%) | Group B n (%) | p-value |
| --- | --- | --- | --- |
| Multiparous | 25 (43.86%) | 26 (45.61%) | 0.8512 |
| Primiparous | 32 (56.14%) | 31 (54.39%) |  |

**Figure 1.**
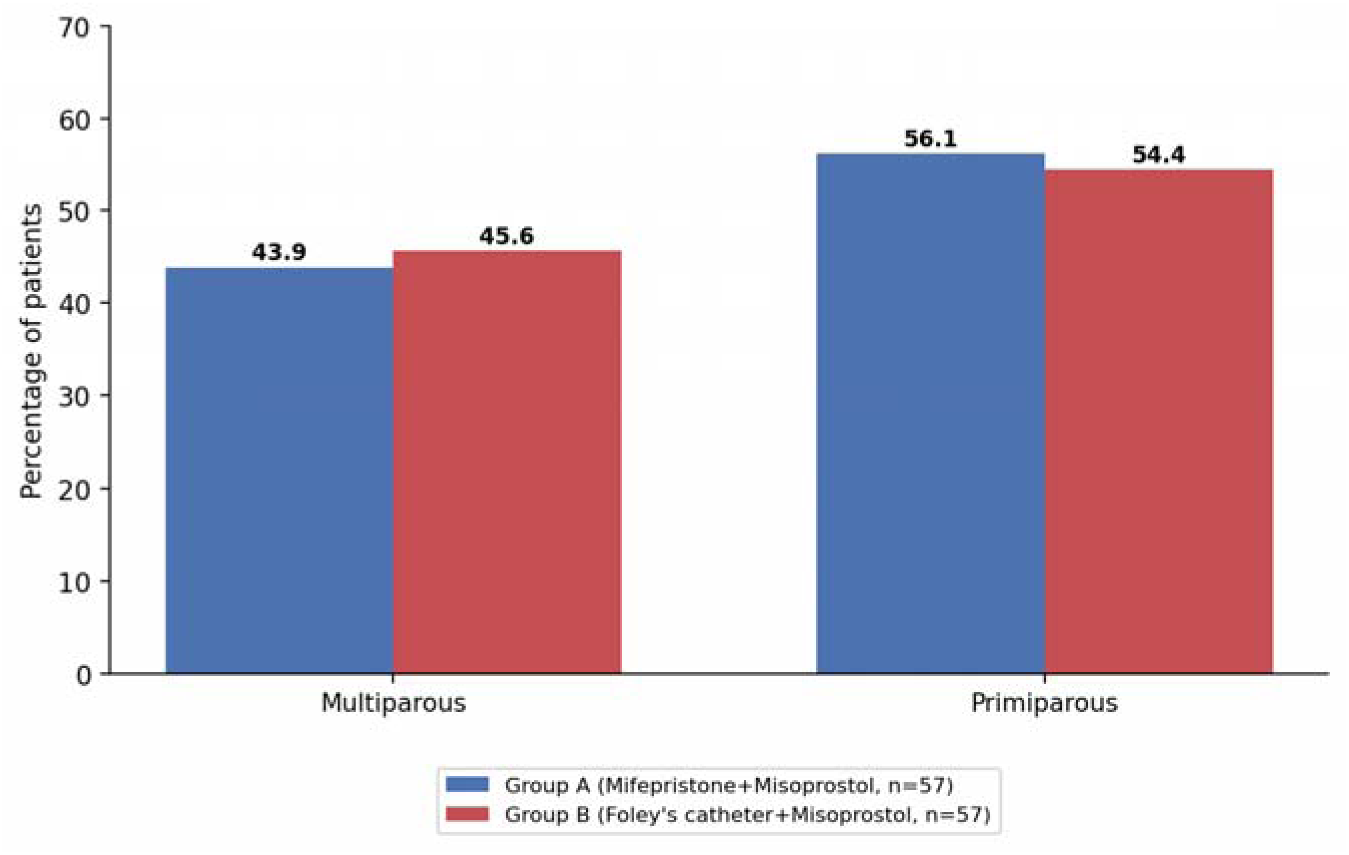
Parity-wise Distribution of Study Population (%)

**Table 4.**
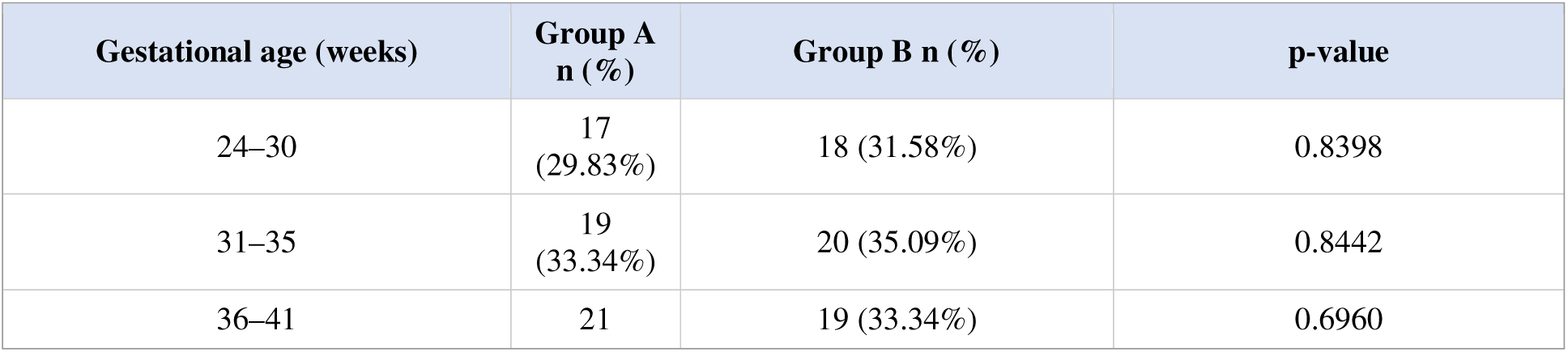
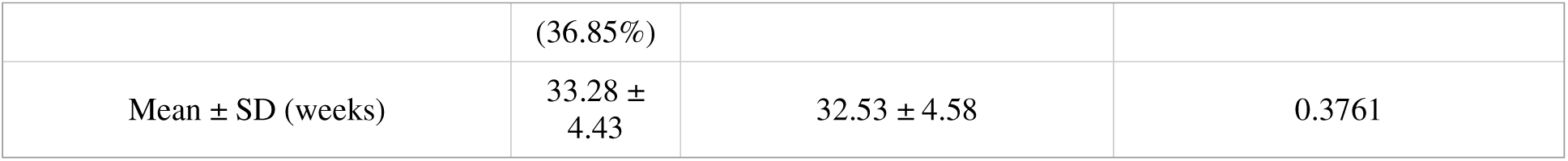
Distribution of study subjects according to gestational age.

**Figure 2.**
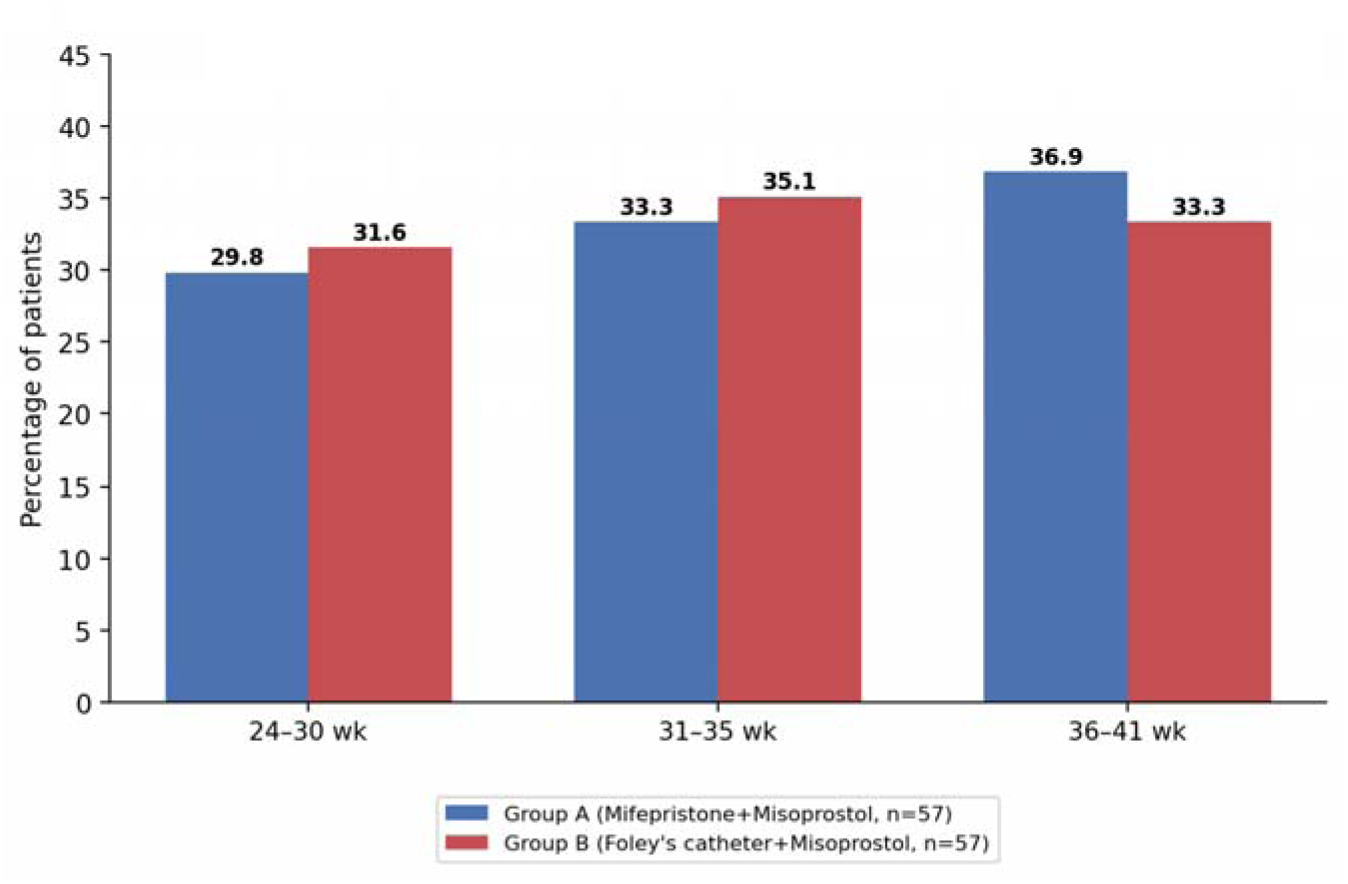
Gestational Age Distribution of Study Population (%)

Baseline age, parity, and gestational age distributions were statistically comparable between the two groups (all p>0.05), confirming successful group balance despite alternate (non-blinded) allocation.

### Cervical response to induction

**Table 5.** Pre-induction Bishop score (0 hour)

| Bishop score | Group A n (%) | Group B n (%) |
| --- | --- | --- |
| 0-3 | 36 (63.15%) | 34 (59.6%) |
| 3-6 | 21 (36.84%) | 23 (40.4%) |
| Mean ± SD | 2.94 ± 1.06 | 2.96 ± 1.13 ($p=0.932$) |

**Table 6.** Post-induction Bishop score (after 24 hours)

| Bishop score | Group A n (%) | Group B n (%) |
| --- | --- | --- |
| <6 | 8 (14.03%) | 12 (21.05%) |
| 6-10 | 46 (80.70%) | 44 (77.19%) |
| >10 | 3 (5.26%) | 1 (1.75%) |
| Mean ± SD | 7.39 ± 2.07 | 6.37 ± 1.89 ($p=0.007$) |

**Figure 3.**
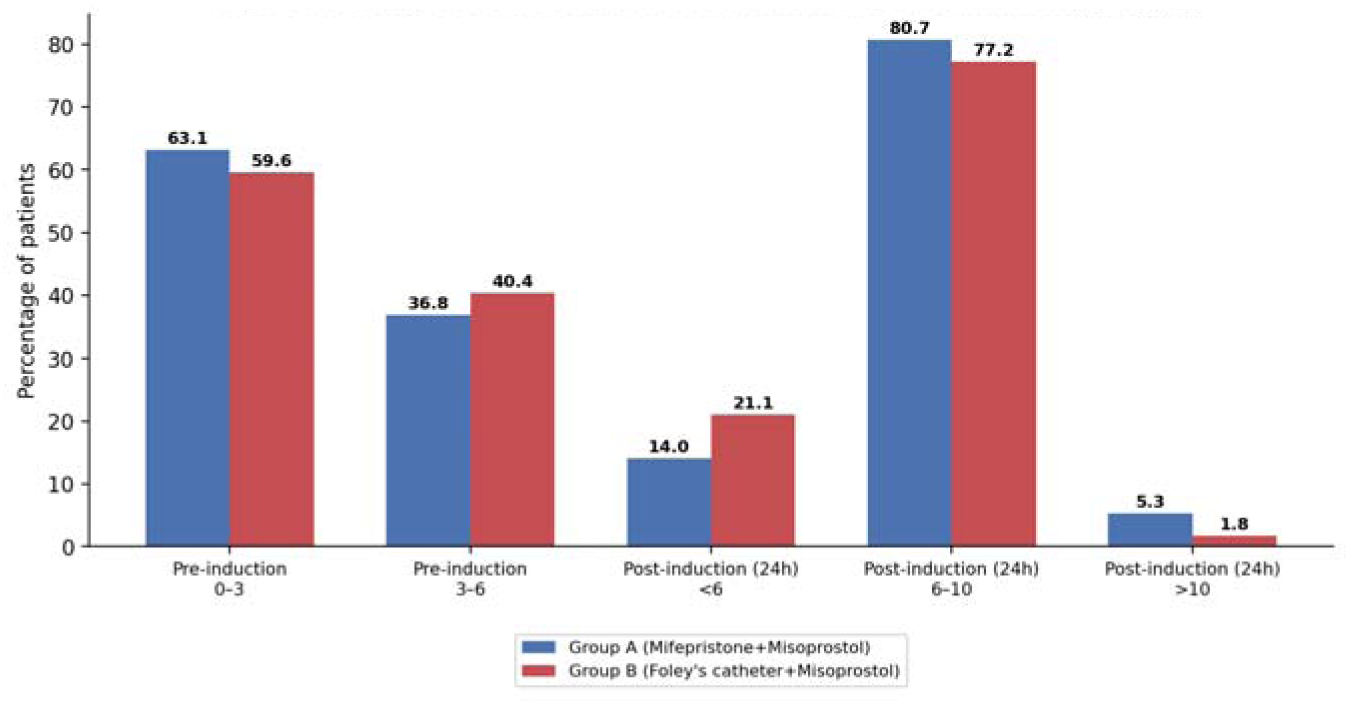
Bishop Score Distribution: Pre-induction vs. Post-induction (24h) (%)

Both groups showed marked improvement in cervical favourability by 24 hours, but the increase in Bishop score was significantly greater in the mifepristone group (p=0.007), consistent with its progesterone-antagonist priming action on the cervix and myometrium.

### Induction-to-delivery interval

**Table 7.** Induction-to-delivery interval according to gestational age (hours, mean ± SD)

|  | Group A | Group B | p-value |
| --- | --- | --- | --- |
| Overall | 25.43 ± 6.84 | 29.26 ± 5.54 | 0.0014 |
| 24–30 weeks | 27.47 ± 7.48 | 32.44 ± 5.45 | 0.0010 |
| 31–35 weeks | 24.37 ± 5.43 | 27.95 ± 5.21 | 0.0425 |
| 36–41 weeks | 24.71 ± 6.99 | 27.63 ± 5.06 | 0.1421 |

**Figure 4.**
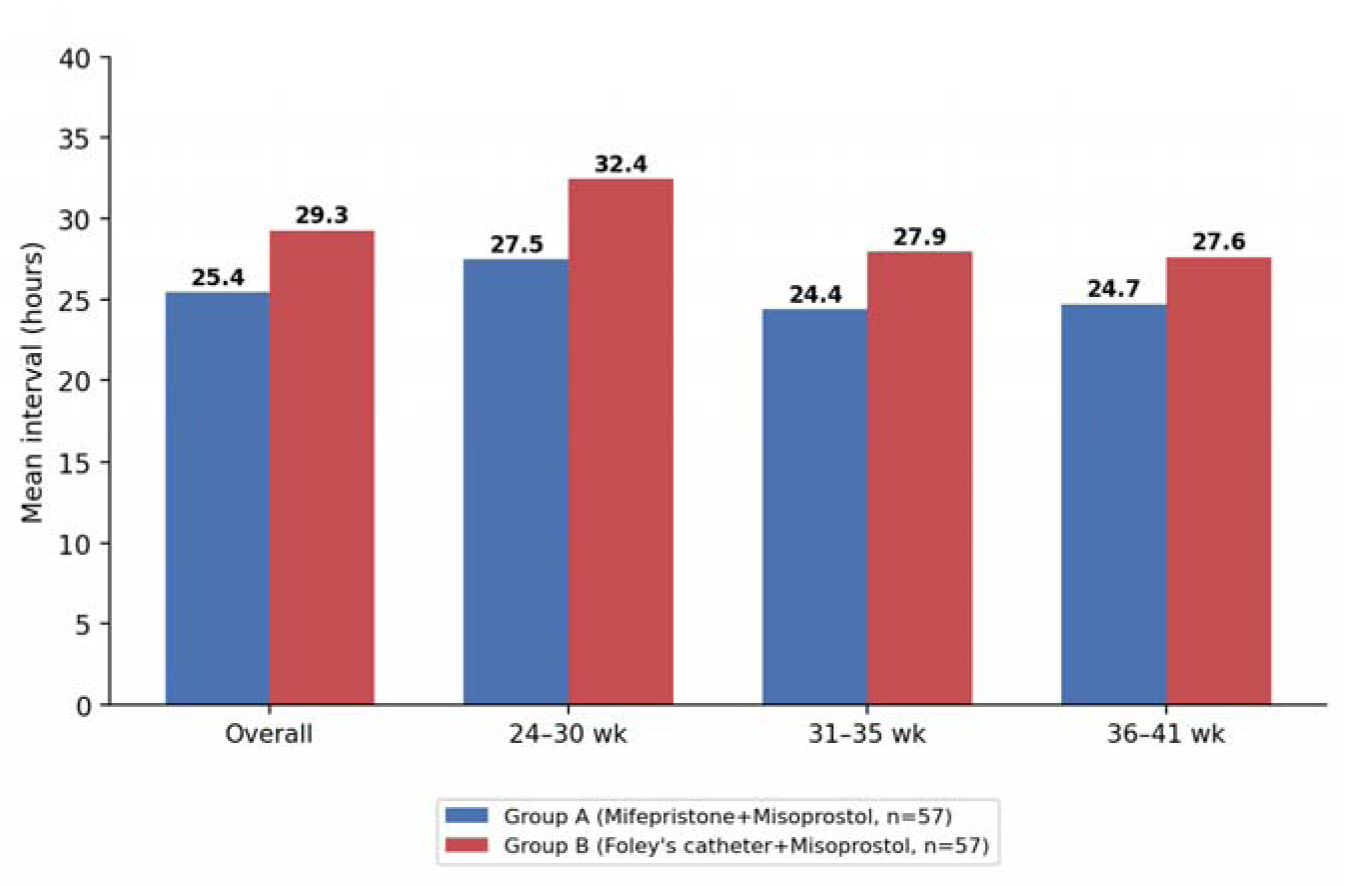
Mean Induction-to-Delivery Interval by Gestational Age (hours)

**Table 8.** Induction-to-delivery interval by pre-induction Bishop score (hours, mean ± SD)

| Bishop score | Group A | Group B | p-value |
| --- | --- | --- | --- |
| 0–3 | 25.74 ± 6.09 | 32.37 ± 4.65 | <0.0001 |
| 3–6 | 21.85 ± 5.85 | 25.96 ± 4.27 | 0.0045 |

**Figure 5.**
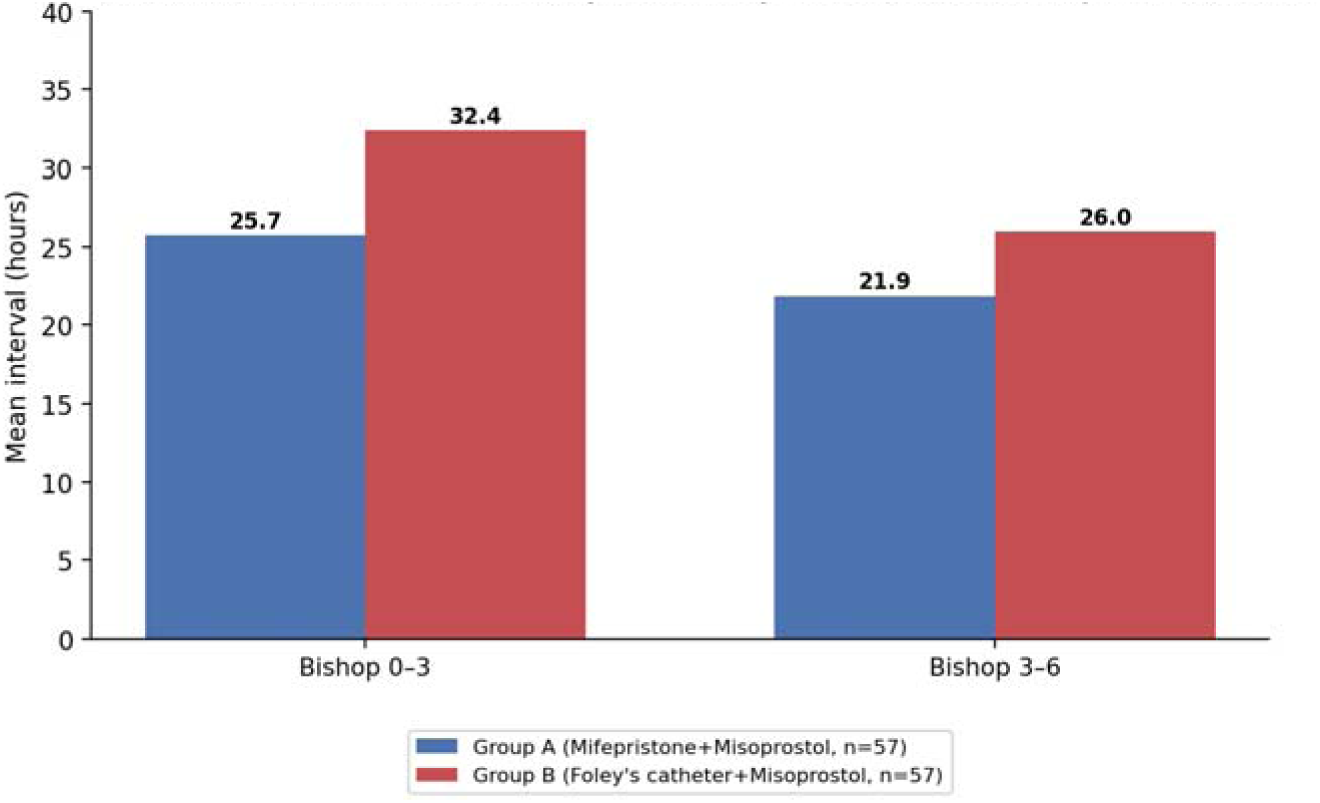
Mean Induction-to-Delivery Interval by Pre-induction Bishop Score (hours)

The induction-to-delivery interval was significantly shorter in the mifepristone group overall and across gestational-age and Bishop-score strata, and shortened further as gestational age advanced and as the pre-induction Bishop score improved, in both groups.

### Misoprostol requirement and oxytocin augmentation

**Table 9.** Total misoprostol dose requirement by gestational age (mcg, median [range])

| Gestational age | Group A | Group B | p-value |
| --- | --- | --- | --- |
| 24–30 weeks | 200 (0–400) | 200 (0–400) | 0.1950 |
| 31–35 weeks | 100 (0–200) | 100 (0–200) | 0.3960 |
| 36–41 weeks | 0 (0–200) | 50 (0–200) | 0.5200 |
| Overall median | 50 (0–100) | 100 (50–200) | <0.01 |

**Table 10.**
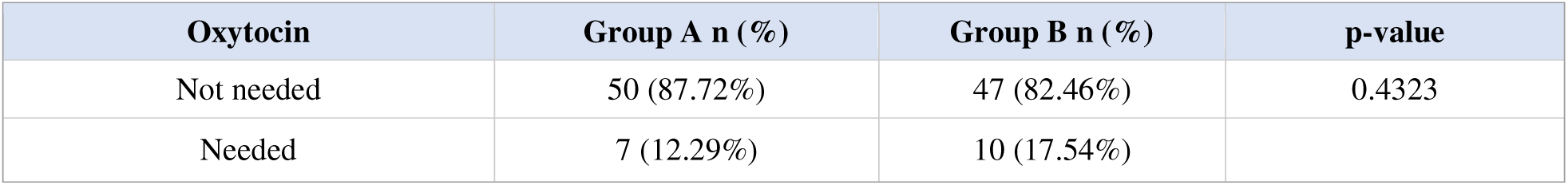
Need for oxytocin augmentation of labour.

| Oxytocin | Group A n (%) | Group B n (%) | p-value |
| --- | --- | --- | --- |
| Not needed | 50 (87.72%) | 47 (82.46%) | 0.4323 |
| Needed | 7 (12.29%) | 10 (17.54%) |  |

**Figure 6.**
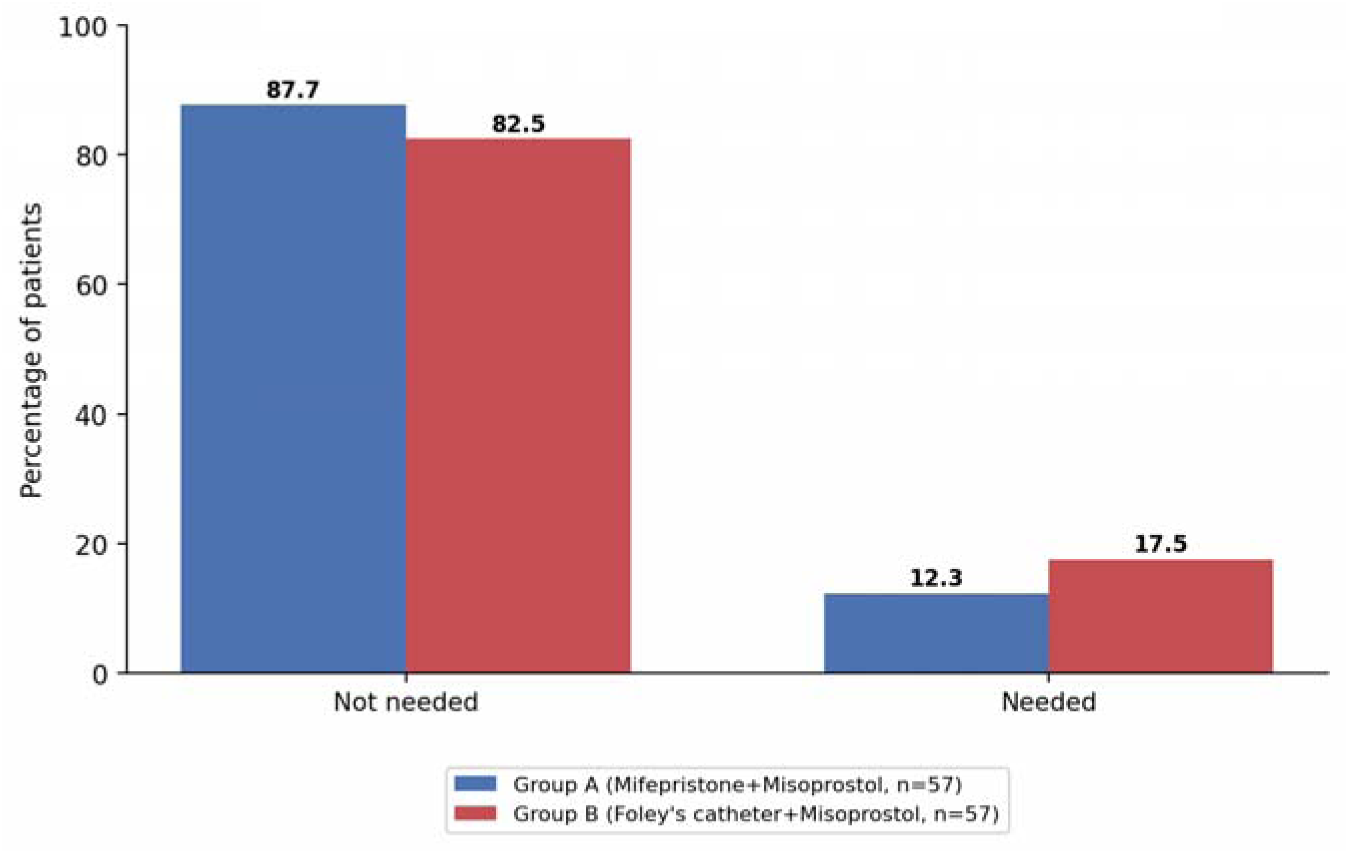
Need for Oxytocin Augmentation of Labour (%)

The total misoprostol dose required was significantly lower in the mifepristone group (median 50 mcg vs. 100 mcg; p<0.01), while the requirement for oxytocin augmentation did not differ significantly between groups.

### Delivery outcomes and maternal complications

**Table 11.** Mode of delivery.

| Mode of delivery | Group A n (%) | Group B n (%) | p-value |
| --- | --- | --- | --- |
| LSCS (caesarean) | 2 (3.51%) | 1 (1.75%) | 1.00 |
| NVD (vaginal) | 55 (96.5%) | 56 (98.24%) |  |

**Figure 7.**
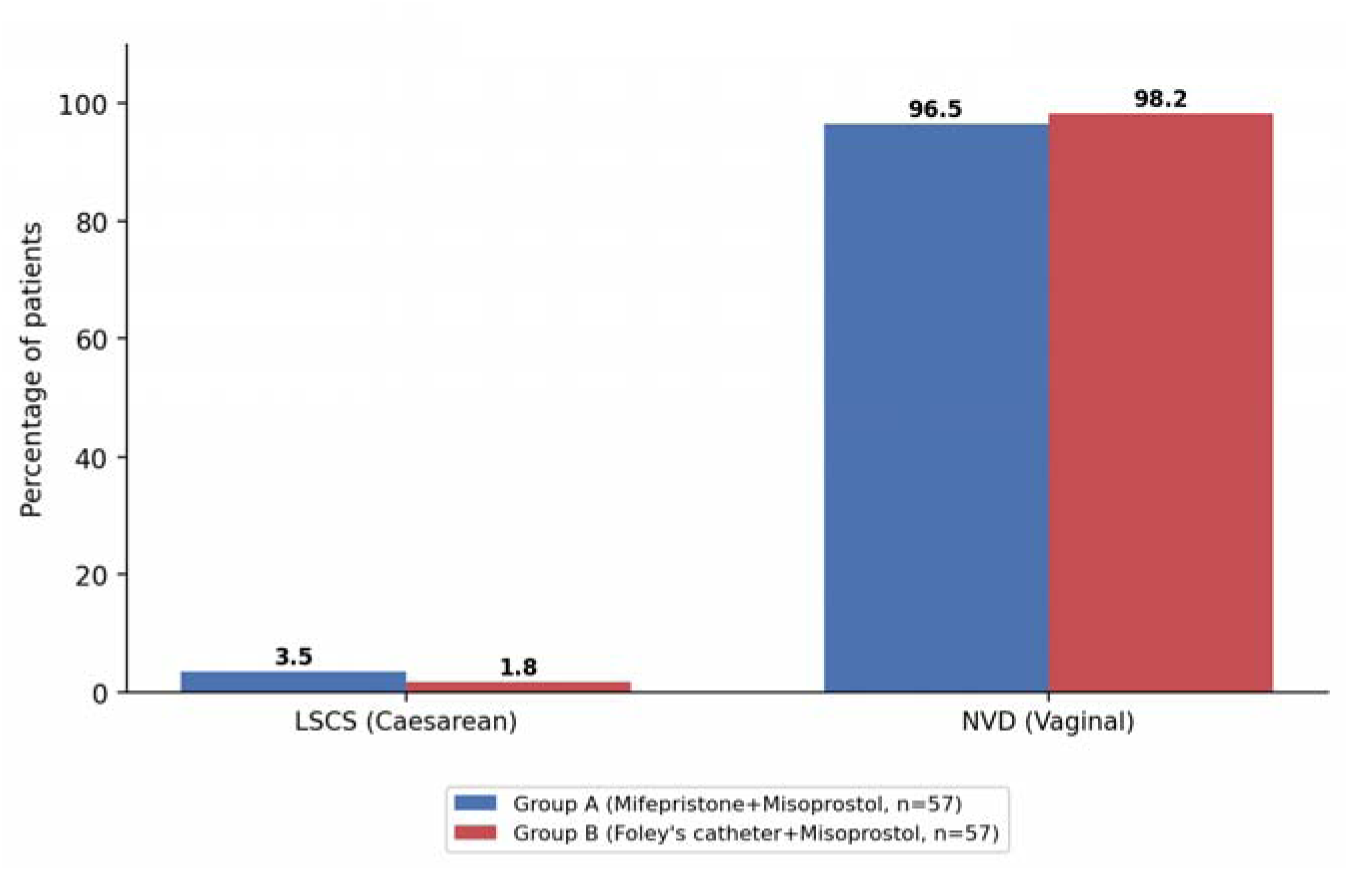
Mode of Delivery (%)

**Table 12.** Amount of blood loss.

| Blood loss (mL) | Group A n (%) | Group B n (%) | p-value |
| --- | --- | --- | --- |
| >500 | 2 (3.51%) | 3 (5.27%) | 0.6489 |
| ≤500 | 55 (96.5%) | 54 (94.74%) |  |

**Figure 8.**
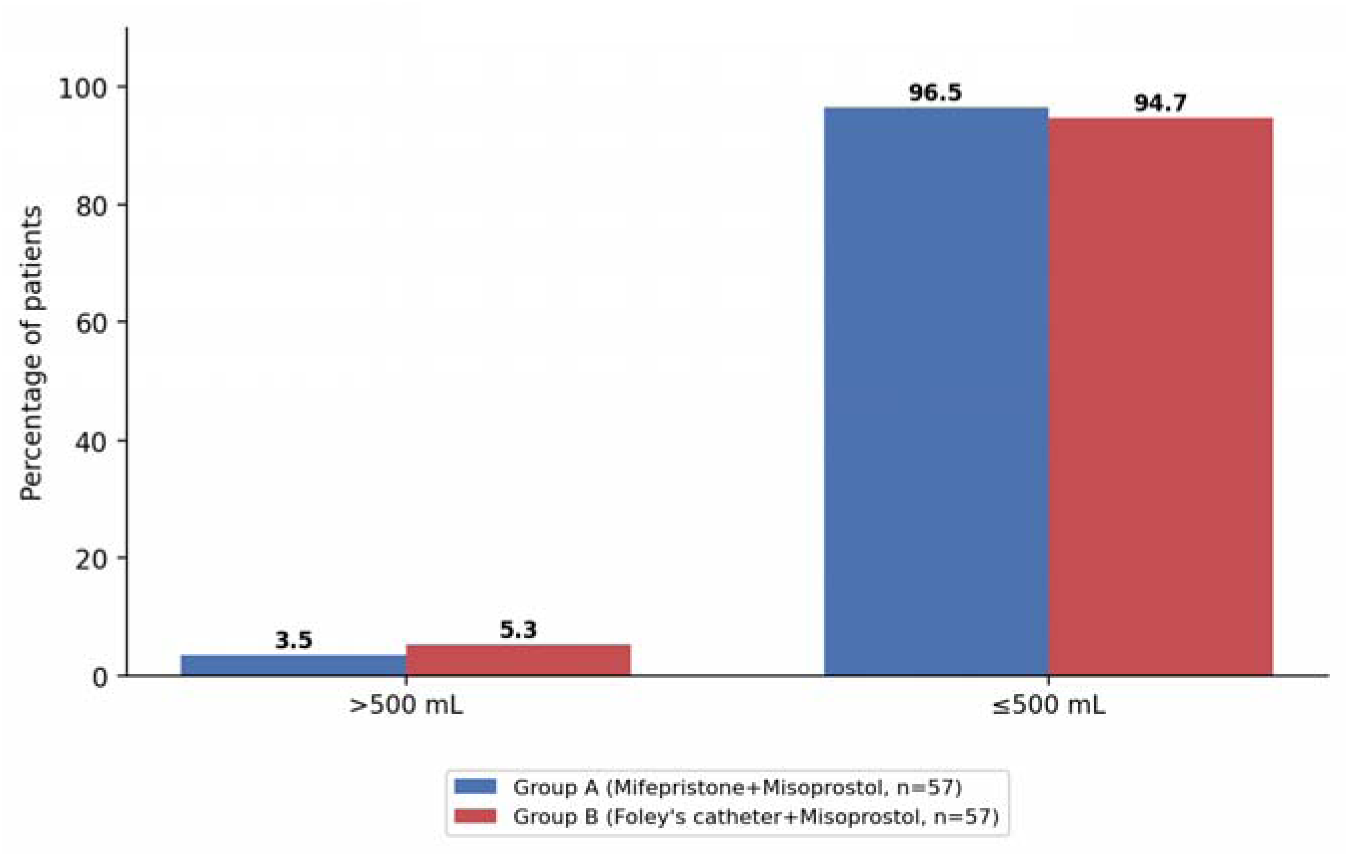
Amount of Blood Loss (%)

**Figure 12.**
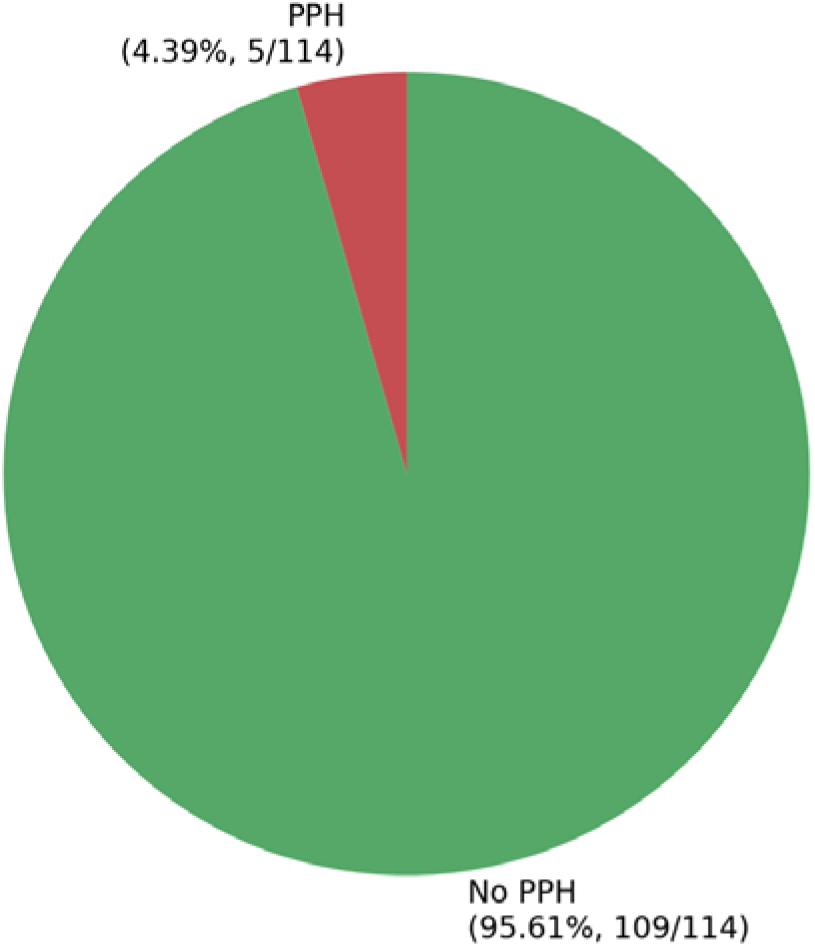
Overall Incidence of Postpartum Haemorrhage (Combined, N=ll4)

**Table 13.**
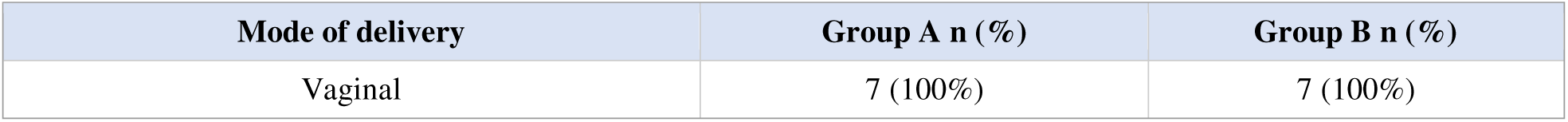
Delivery outcome in patients with previous caesarean section (N=7)

| Mode of delivery | Group A n (%) | Group B n (%) |
| --- | --- | --- |
| Vaginal | 7 (100%) | 7 (100%) |

**Table 14.** Patients in Group A delivered with mifepristone alone (n=57)

| Additional misoprostol required? | n | % |
| --- | --- | --- |
| No (mifepristone alone sufficient) | 24 | 42.11% |
| Yes (misoprostol required) | 33 | 57.9% |

**Figure 13.**
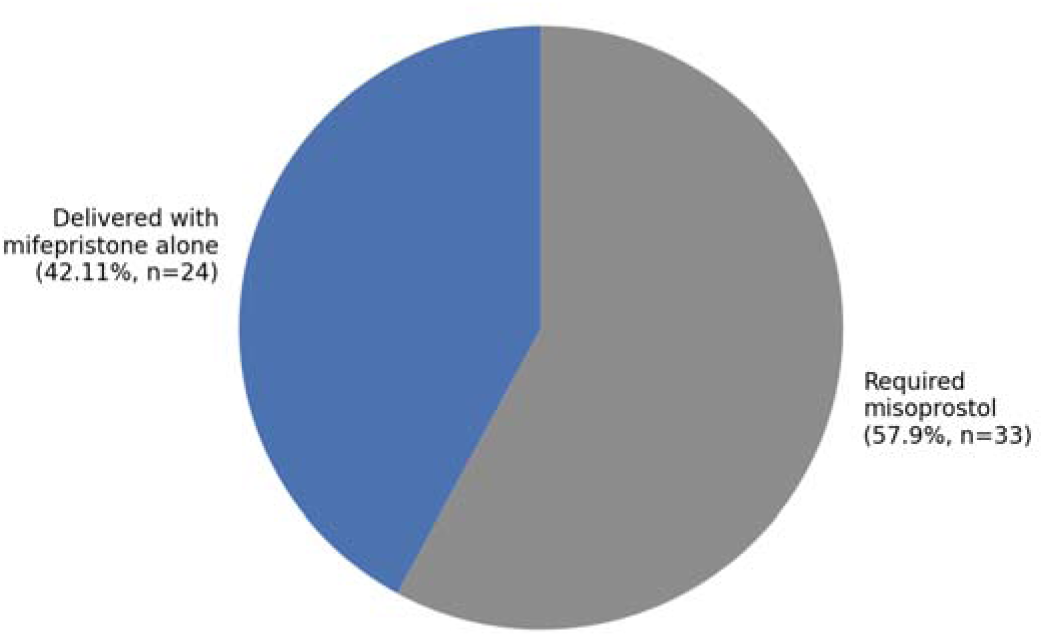
Proportion of Group A Patients Delivered with Mifepristone Alone (n=57)

**Table 15.**
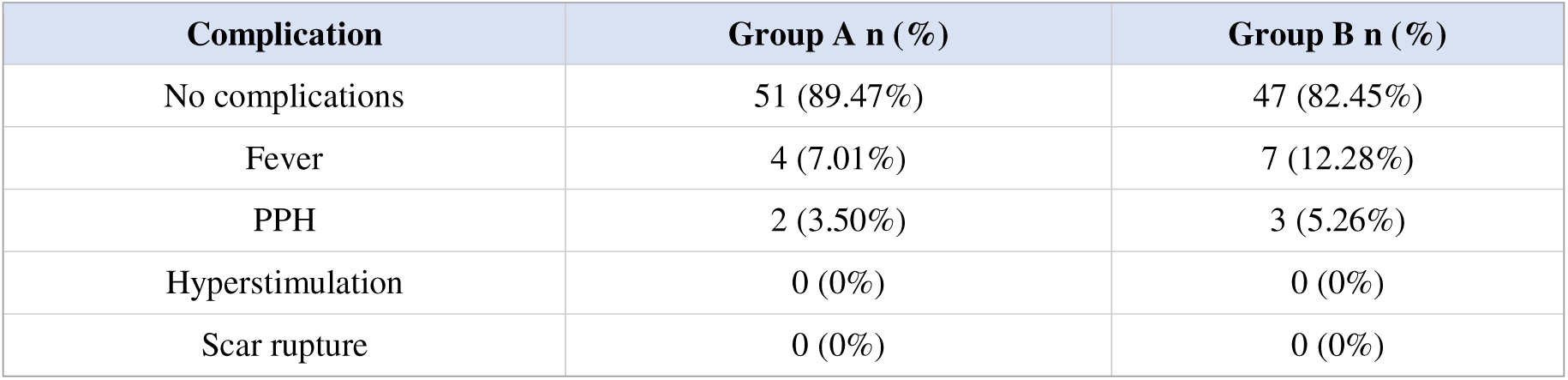
Maternal complications.

**Figure 9.**
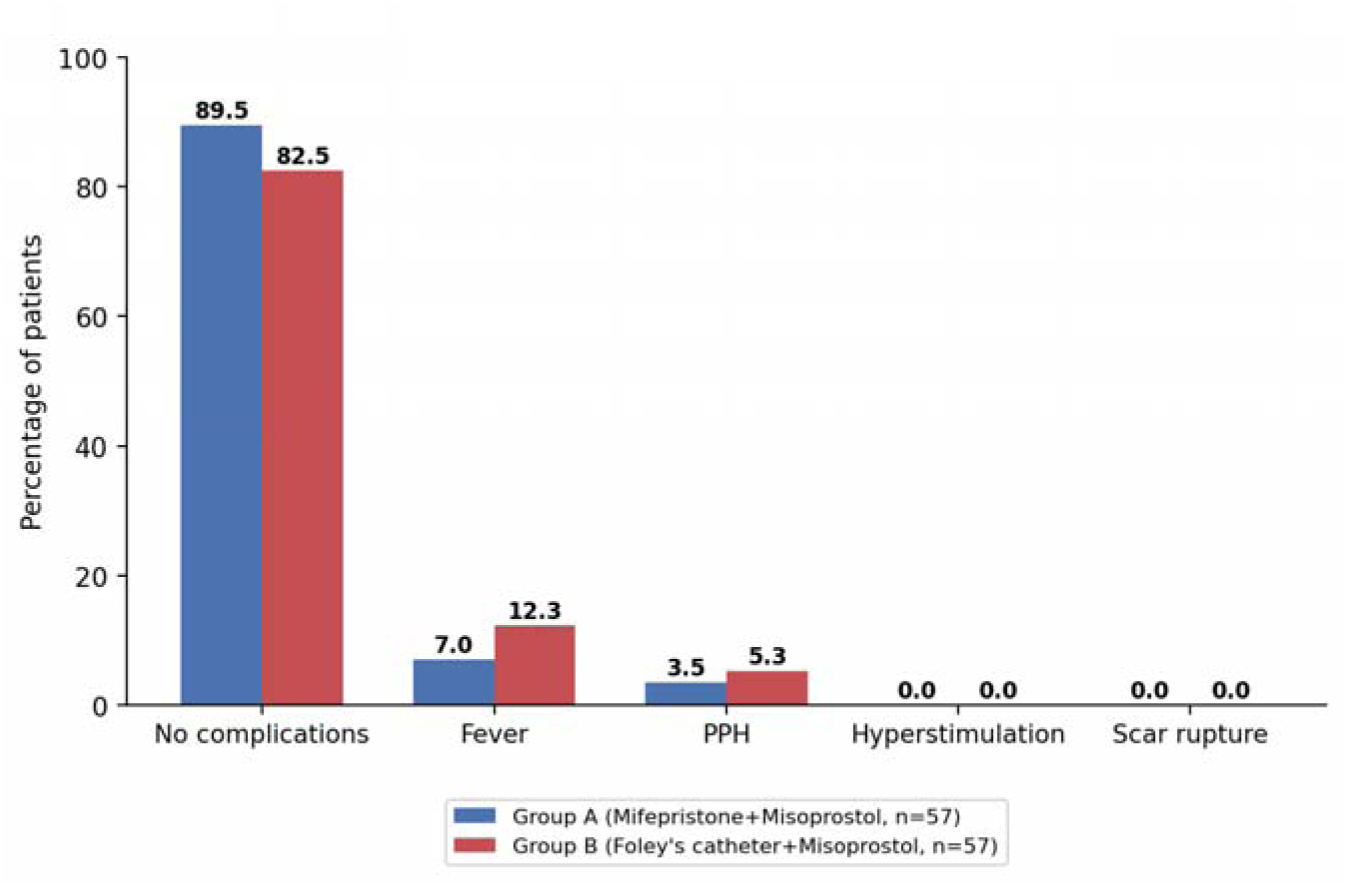
Maternal Complications (%)

The overall complication rate, mode of delivery, blood loss, and rate of PPH were statistically comparable between the two groups (all p>0.05). No cases of uterine hyperstimulation or scar rupture occurred in either group, including among the seven women with a prior caesarean delivery, all of whom delivered vaginally.

### Pain and patient satisfaction

**Table 16.**
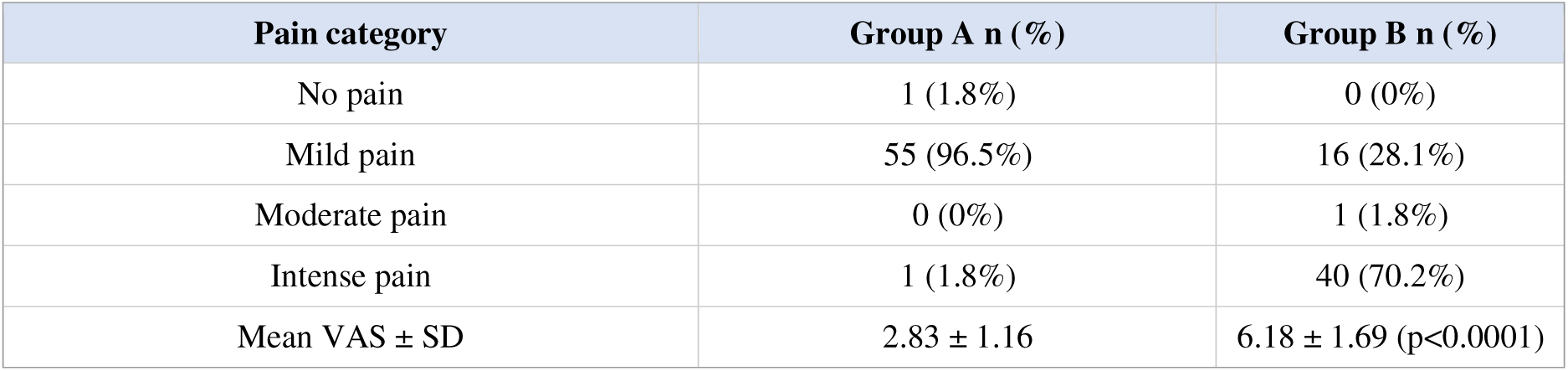
Pain score (visual analogue scale, VAS) during cervical ripening.

| Pain category | Group A n (%) | Group B n (%) |
| --- | --- | --- |
| No pain | 1 (1.8%) | 0 (0%) |
| Mild pain | 55 (96.5%) | 16 (28.1%) |
| Moderate pain | 0 (0%) | 1 (1.8%) |
| Intense pain | 1 (1.8%) | 40 (70.2%) |
| Mean VAS ± SD | 2.83 ± 1.16 | 6.18 ± 1.69 ($p < 0.0001$) |

**Figure 10.**
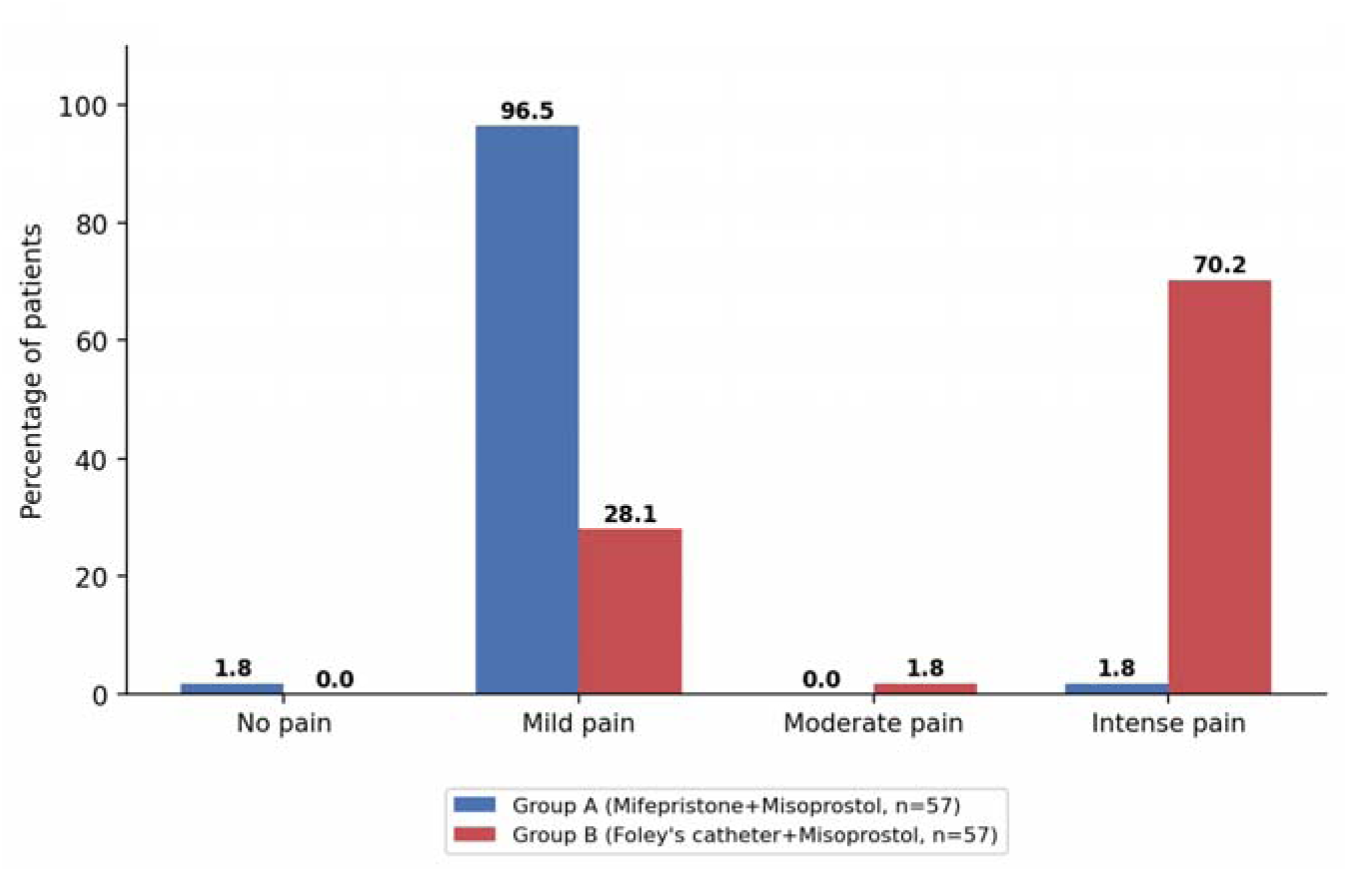
Pain Score (VAS) Distribution during Cervical Ripening (%)

**Table 17.** Patient satisfaction with the induction method.

| Satisfaction | Group A n (%) | Group B n (%) | p-value |
| --- | --- | --- | --- |
| No | 2 (3.51%) | 5 (8.78%) | 0.2439 |
| Yes | 55 (96.5%) | 52 (91.23%) |  |

**Figure 11.**
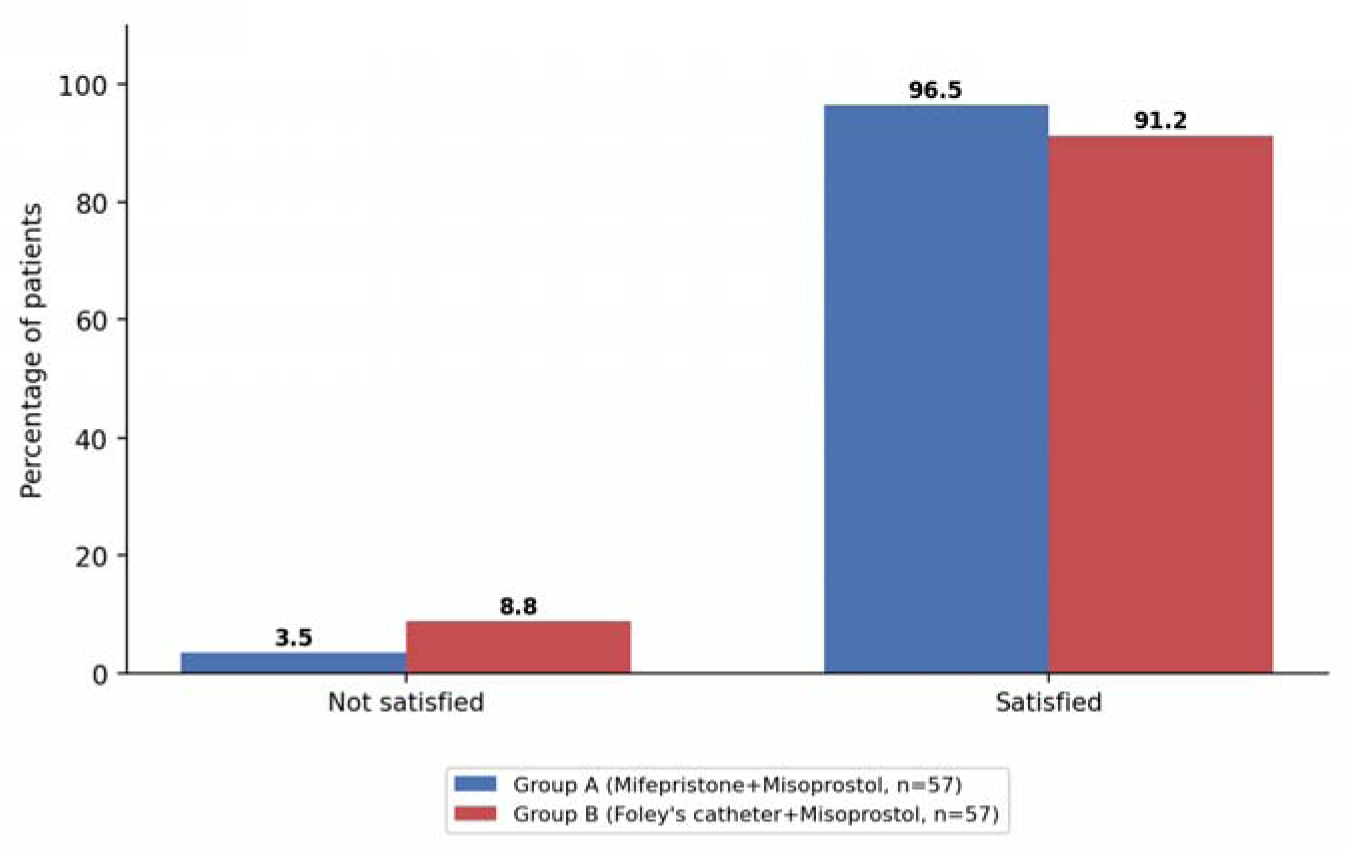
Patient Satisfaction with Induction Method (%)

Pain during cervical ripening was significantly lower in the mifepristone group (mean VAS 2.83±1.16) than in the Foley’s catheter group (mean VAS 6.18±1.69; p<0.0001), while overall patient satisfaction, though numerically higher with mifepristone, did not differ significantly between groups.

## Discussion

In the present study, both mifepristone-misoprostol and Foley’s catheter-misoprostol regimens achieved a favourable cervix and vaginal delivery in the large majority of women with IUFD beyond 24 weeks of gestation, with an overall vaginal delivery rate exceeding 96% in both groups. This is consistent with prior comparative studies of these two regimens across other obstetric populations, which have similarly reported high rates of successful vaginal delivery with both methods. ^[1,3,4]^

The significantly shorter induction-to-delivery interval observed with mifepristone priming in this study (25.43±6.84 hours vs. 29.26±5.54 hours) is consistent with the findings of Väyrynen et al., who compared misoprostol alone against mifepristone plus misoprostol for induction following intrauterine fetal death and similarly reported a shorter interval with mifepristone priming. ^[1]^ Wing et al. also reported that mifepristone priming shortened the interval to active labour compared with placebo in preinduction cervical ripening. ^[4]^ Lata et al. and Yelikar et al. reported comparable reductions in induction-to-delivery interval with mifepristone priming in term pregnancies, supporting the generalisability of this effect across gestational contexts. ^[2,5]^

The lower total misoprostol requirement in the mifepristone group in this study mirrors the mechanism proposed by several authors: mifepristone’s antiprogestin action increases myometrial gap-junction formation and oxytocin-receptor expression, priming the uterus for a more efficient prostaglandin response and thereby reducing the cumulative prostaglandin dose needed to achieve active labour. ^[1,2]^ This is clinically relevant in resource-limited settings, where minimising drug requirement and repeat dosing reduces both cost and nursing workload.

The markedly lower pain scores observed with mifepristone priming in this study are consistent with the mechanical nature of Foley’s catheter placement and inflation, which is inherently more uncomfortable than oral tablet administration; Väyrynen et al. and Fonseca and Sah similarly reported greater patient discomfort with Foley’s catheter compared with pharmacological priming regimens. ^[1,6]^ Despite this difference, overall patient satisfaction was high and statistically comparable between groups in the present study, suggesting that successful, timely vaginal delivery may be a more important determinant of overall satisfaction than procedural discomfort alone.

Importantly, maternal complication rates — including postpartum haemorrhage, fever, uterine hyperstimulation, and scar rupture — did not differ significantly between the two groups, and all seven women with a prior caesarean delivery in this cohort delivered vaginally without scar complications, supporting the safety of both regimens, including in women with a uterine scar, findings consistent with prior reports on Foley’s catheter safety in scarred uteri. ^[3]^

## Strengths and limitations

This study’s principal strengths are its prospective design, adequately matched baseline characteristics between groups, and comprehensive assessment of both efficacy (induction-to-delivery interval, misoprostol requirement) and patient-centred outcomes (pain, satisfaction). Limitations include the relatively modest sample size, which may limit precision for less common outcomes such as PPH and uterine rupture; the absence of a third, no-treatment or alternative-agent control group; and the use of alternate (rather than computer-generated random) allocation, which, while standard in many resource-limited obstetric units, does not eliminate all potential allocation bias.

## Conclusions

In women with intrauterine fetal death beyond 24 weeks of gestation and an unfavourable cervix, both sequential mifepristone-misoprostol and intracervical Foley’s catheter-misoprostol regimens were safe, effective, and acceptable methods for cervical ripening and induction of labour, with vaginal delivery achieved in more than 96% of women in both groups. Mifepristone priming was associated with a significantly shorter induction-to-delivery interval, a lower total misoprostol requirement, a greater improvement in Bishop score at 24 hours, and substantially less procedural pain, and may be preferred as a first-line regimen where mifepristone is available. Foley’s catheter with misoprostol remains a safe, simple, and low-cost alternative, particularly relevant where mifepristone access is limited, though it was associated with greater patient discomfort during cervical ripening. Larger, multicentric studies are recommended to confirm these findings and to further characterise safety in women with a prior uterine scar.

## Supporting information

Title Page

## Data Availability

GRMC gwalior

